# DrugSet: A validated R Shiny application for reproducible drug codelist construction from ATC classification to CPRD Aurum prodcodes

**DOI:** 10.64898/2026.07.08.26357534

**Authors:** Vjola Hoxhaj, Catherine Fry, Denise E Morris, Taylor Aurelius, Samantha Lane, Miriam Sturkenboom, Constanza L. Andaur Navarro

## Abstract

**KEY POINTS:**

- Constructing semantically harmonized drug codelists for pharmacoepidemiologic research across data sources is often manual, time-consuming, and prone to inconsistencies as many datasources but not all (e.g. CPRD Aurum) use the ATC coding system.
- DrugSet is a validated open-source R Shiny application that integrates ATC-based hierarchical code selection, string-based matching on pre-specified variables mapping to CPRD Aurum prodcodes, and codelist export within a single workflow operating directly on native vocabulary files.
- DrugSet demonstrated high accuracy for both ATC codelist generation and prodcodes mapping, and reduced the time required for codelist construction compared to manual approaches.

**PLAIN LANGUAGE SUMMARY:** Health research focused on use and safety of medicines depends on accurate lists of drug codes, called codelists, to identify which medicines the patients received. Building these lists is currently a manual process, which makes it slow, error-prone, and difficult to reproduce across studies.

We developed DrugSet, an open-source tool that helps researchers build drug codelists in a structured and consistent way. The tool uses the international drug classification system (ATC: Anatomical Therapeutic Chemical) to identify drug groups of interest and automatically links these to the specific codes used in the largest United Kingdom primary care data source (CPRD: Clinical Practice Research Datalink Aurum). By leveraging the route-of-administration information already embedded in the ATC hierarchy, it also distinguishes, for example, topical from systemic formulations. We tested DrugSet on three drug types, metformin, beta-blockers, and topical salicylic acid and found that it produced results matching reference standards, completing the task in minutes rather than hours. DrugSet is freely available to support transparent, reproducible for pharmacoepidemiological research.

**Objectives:** To present DrugSet, a validated R Shiny application supporting the construction medicinal products codelists based on the Anatomical Therapeutic Chemical (ATC) system and their mapping to Clinical Practice Research Datalink (CPRD) Aurum prodcodes within a single interactive workflow.

**Materials and Methods:** DrugSet comprises four modules: data preparation, ATC-based hierarchical code selection, string-based CPRD Aurum prodcodes mapping, and codelist export. Validation was conducted against World Health Organization (WHO) ATC reference codelists and manually curated prodcodes mappings across three drug classes: metformin, beta-blocking agents, and topical salicylic acid. Sensitivity, specificity, and Positive Predictive Values (PPV) were calculated for ATC codelist generation. Agreement proportions (overlapping against total identified codes) were calculated for prodcodes mapping. Time needed for codelist construction using DrugSet was recorded and compared to manual approaches.

**Results:** DrugSet’s ATC codelist generation against WHO manual reference achieved 100% sensitivity, specificity, and PPV across all medicinal products. Prodcodes mapping agreement ranged from 89.2% to 98.3% with discrepancies due to missing data in the prodcodes input vocabulary. DrugSet completed codelist construction in 9 minutes compared to 3 hours and 10 minutes manually, across all medicinal products classes.

**Discussion:** DrugSet provides a unified workflow that runs directly on ATC and source CPRD Aurum vocabulary files. The reduction in codelist construction time and export of the generated codelists supports reproducibility in pharmacoepidemiologic studies where codelist creation can represent a significant proportion of study setup time.

**Conclusion:** DrugSet is an open-source, validated tool that improves accuracy, and efficiency of codelist construction for medicinal products based on ATC codes towards CPRD Aurum prodcodes.

## INTRODUCTION

Real-world data (RWD) have been important in pharmacoepidemiology to generate evidence on the use, effectiveness, and safety of medicinal products since the last century and are recognized as important source of information by regulatory agencies in North America and Europe.^1–4^ However, because RWD are collected for health care delivery or payment rather than research, medicinal products are captured differently across sources through dispensing, prescriptions, or administrations. To estimate exposure to medicinal products, accurate retrieval of such information relies on codelists, a curated set of codes that operationalizes how medicinal products of relevance for a study can be identified in a data source.

In RWD sources, medicinal products are often recorded via a national product code used in pharmacy dispensing records and for printing prescriptions, which in some sources is mapped to the hierarchical Anatomical Therapeutic Chemical (ATC) classification, but in others, particularly prescription-based systems, may not be linked to ATC at all. The ATC classification, maintained by the World Health Organization (WHO), is widely used internationally and organizes medicinal products hierarchically across 5 levels: the 1st level reflects the organ or body system acted on, the 2nd the therapeutic subgroup, the 3rd and 4th further therapeutic or chemical subgroups, and the 5th the chemical substance.^5,6^ A single medicinal product may map to multiple ATC codes depending on indication (e.g., acetylsalicylic acid for pain versus antiplatelet therapy), while a single ATC code may conversely cover distinct products differing in formulation, strength, or route (e.g., mesalazine oral and rectal formulations). Other data sources use product-level identifiers with no link to any standardized classification: the UK Clinical Practice Research Datalink (CPRD) Aurum, for instance, assigns prodcodes at the point of prescribing based on a primary care system specific IDs with no inherent clinical or chemical meaning.^7,8^ In the US, RxNorm sits between these extremes, providing normalized product names, cross-vocabulary links, and a mapping to ATC.^9^

The coexistence of multiple, structurally different coding systems for medicinal product means researchers must manually map higher-level classifications like ATC codes to data source-specific dictionaries, a process that is often manual, iterative (need for validation and refinement of codes), time-consuming, and complicated by periodic updates. For example the WHO ATC/DDD (Defined Daily Dose) is updated annually by adding new ATC codes, classification changes for existing medicinal products, correction of substance names and so on, requiring researchers to confirm alignment between codelists and the version used in the data source.^10^

Several tools and standards support codelist creation, but each is tailored to a specific data model or dictionary. On the regulatory side, the Identification of Medicinal Products (IDMP) standards and the European Medicines Agency’s Substance, Products, Organizations, Referential (SPOR) framework aim to harmonize medicinal product information across regulatory systems, but they are not yet uniformly implemented in routinely collected data used for observational research, meaning source-specific dictionaries remain widely used especially in legacy data.^11,12^ Among codelists tools, OpenCodelists (OpenSAFELY) supports creation and sharing of medicinal product codelists based on National Health Service (NHS) terminologies such as dm+d (UK standard medicinal product/device coding system) and SNOMED CT (clinical terminology system).^13,14^ The wardle/dmd tool enables mapping between ATC or British National Formulary (BNF) codes to dm+d product concepts.^15^ Within federated research networks such as DARWIN EU, the CodelistGenerator supports medicinal product codelists derivation from Observational Medical Outcomes Partnership (OMOP) standardized dictionaries based on RxNorm.^16^ For CPRD Aurum specifically, Graul et al. described a semi-automated, clinician-reviewed methodology for generating standardized and reproducible codelists for medicinal products, by searching multiple medical product dictionary variables for chemical and proprietary drug names, alongside the BNF ontology codes, generating prodcodes codelists.^8^ Despite these advances, no existing tool provides a unified workflow for generating ATC-based medicinal product exposures and mapping them to CPRD Aurum prodcodes within a single environment.

To address this gap, we present DrugSet, an R shiny application designed to support generation of codelists for medicinal products based on ATC classification and their mapping to the CPRD Aurum prodcodes. The application integrates data ingestion, preprocessing, hierarchical code selection, and mapping within a single workflow.

## METHODS

### Data input and workflow

DrugSet processes input files stored locally within the Scripts/Dictionaries/ directory (Figure 1). Supported file formats include .csv, .txt, and .xlsx, and files should be labelled starting with prefixes “ATC_” for ATC-datasets and “CPRD_” for CPRD-dataset, allowing multiple files to be loaded and combined.

**Figure 1.**
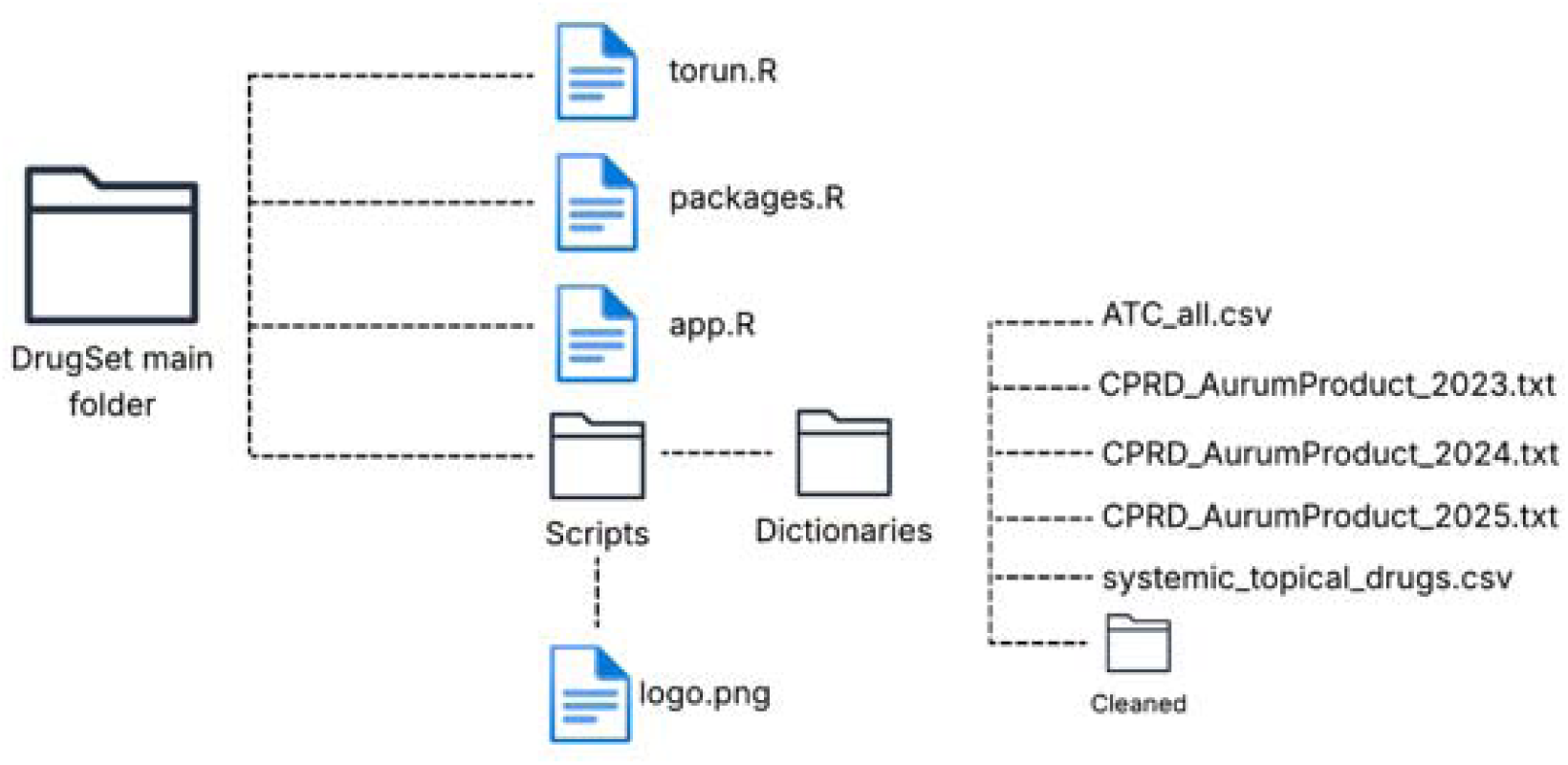
DrugSet folder structure.

For ATC-datasets, users specify the columns corresponding to medicinal product codes and medicinal product names in text boxes in the application interface. For CPRD datasets, required variables include code, EMIS term, product name, substance name, formulation, and administration route. These variables are mapped to standardized column names during preprocessing (Box 1).

**Box 1.** Variable descriptions and example for ATC-based and CPRD-based datasets.

| Column name | Description | Example |
| --- | --- | --- |
| **ATC-based datasets** |  |  |
| **code** | ATC codes identifier | A10BA02 |
| **drug_name** | Name of medicinal product/substance | Metformin |
| **CPRD-based datasets** |  |  |
| **code** | Prodcodes identifier | 644941000033117 |
| **EMIS_term** | Clinical term used in EMIS* | Glucophage 500mg tablets<br>(Merck Serono Ltd) |
| **product_name** | Product name | Glucophage 500mg tablets |
| **substance_name** | Active substance name | Metformin hydrochloride |
| **formulation** | Medicinal product formulation | Tablet |
| **administration_route** | Route of administration | Oral |

\*EMIS: Egton Medical Information Systems, the UK clinical software used in general practice from which data are collected to populate the CPRD Aurum database.

The DrugSet application consists of four modules and three tabs: (1) Prepare and clean input files, (2) Create a medicinal product codelist: ATC-based codelist generation and mapping to prodcodes, and (3) Export full medicinal product codelist. A central working dataset is maintained throughout the workflow and updated iteratively as new entries are generated or modified (Box 2).

**Box 2.** Overview of DrugSet workflow modules.

| **Module** | **Description** | **Input** | **Output** | **Tab** |
| --- | --- | --- | --- | --- |
| **Data preparation** | Standardization and cleaning of input files | Source WHO ATC and CPRD Aurum prodcodes files | Cleaned datasets (ATC_cleaned.csv and CPRD_cleaned.csv) | Prepare and clean input files |
| **ATC codelist generation** | Selection and expansion of ATC codes | ATC cleaned dataset | Working codelist (ATC-based) | Create a medicinal product codelist |
| **Mapping** | Mapping ATC-based entries to prodcodes | Working codelist + CPRD cleaned dataset | Mapped codelist |  |
| **Export** | Output generation | Final working codelist | Exported file | Export full medicinal product codelist |

### Application design and implementation

DrugSet is implemented in R using the Shiny framework, combining a graphical user interface with server-side data processing. The application is organized into four sequential modules corresponding to the workflow steps. The application maintains and updates a central data object (i.e., working codelist) across modules to store selected codes and associated metadata (Figure 2). This working codelist contains the variables of drug_abbreviation, coding_system, code, product_name, label, and drug_concept.

**Figure 2.**
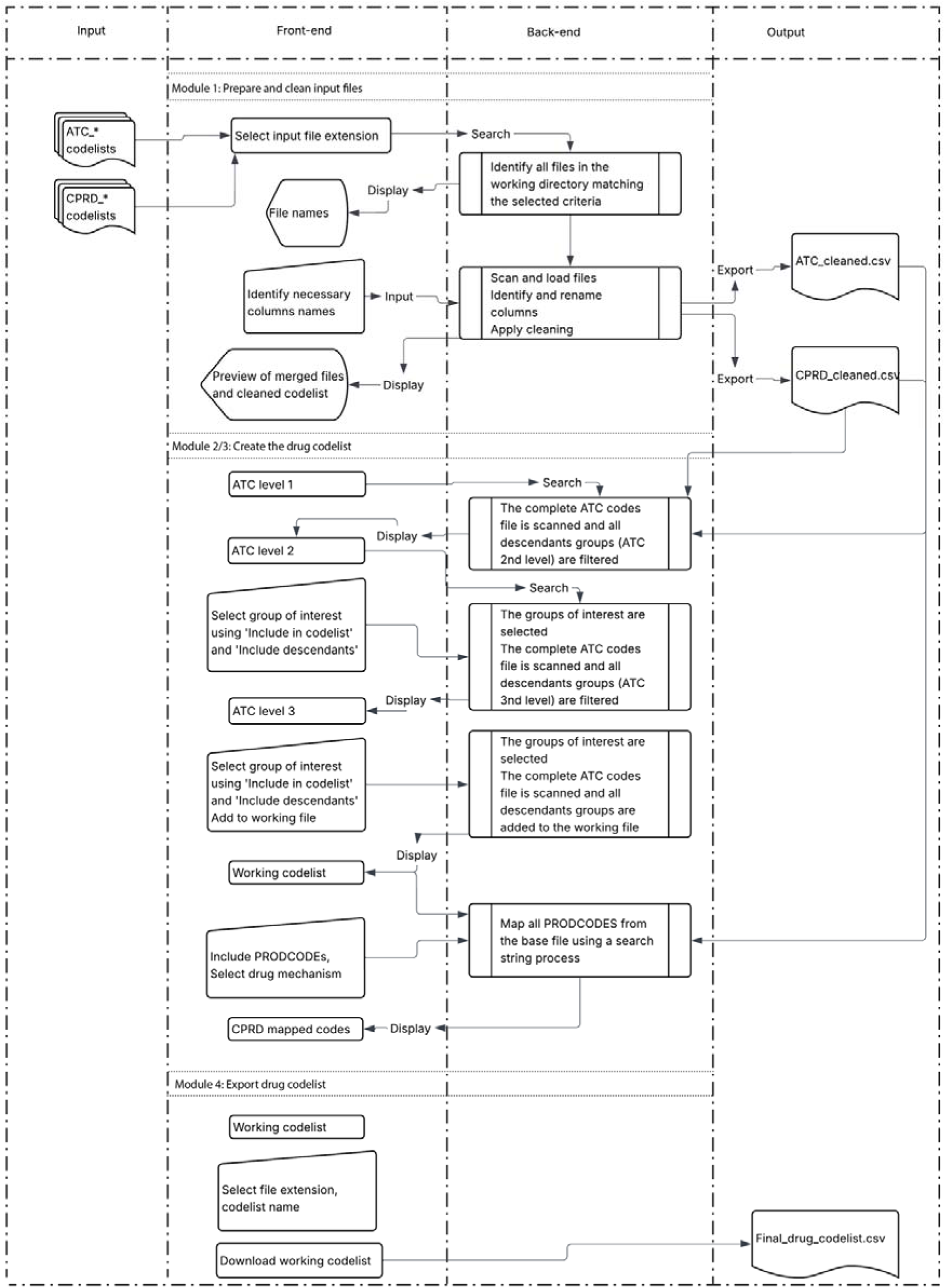
DrugSet workflow.

Two user-defined parameters are required prior to codelist construction (Box 3). The variable drug_abbreviation is used as a grouping identifier and is automatically formatted to include the prefix “DC_”, with whitespace replaced by underscores. The variable drug_concept provides a descriptive label for the medicinal product class. The other parameters that are automatically assigned are coding system as ATC, and label as DC_Proxy to show that this is a medicinal product concept proxy. These parameters are assigned to all generated entries and retained throughout the workflow.

**Box 3.** User-defined parameters for medicinal product codelist construction.

| Column name | Description | Example |
| --- | --- | --- |
| drug_abbreviation | group identifier | DC_Metformin |
| coding_system | Coding system vocabulary | ATC |
| code | ATC code value | A10BA02 |
| product_name | Name of medicinal product/substance | Metformin |
| label | Label for the codelist | DC_Proxy |
| drug_concept | Short description of the medicinal product class | glucose_lowering |

### Data preparation

In the first module, the application processes source ATC and CPRD input files, which are publicly available (ATC) or accessible through a license (CPRD).^7,17^ The application scans the pre-specified directory for files matching the selected extension and naming pattern and merges the identified files into a unified dataset. For this project, the complete ATC codelist (version 2025) from WHO and three .txt input files (2023, 2024 and 2025 version) for CPRD Aurum prodcodes were used. Users define the column names corresponding to required variables, which are then mapped to standardized column names. Preprocessing includes selection and renaming of relevant columns, removal of rows with missing or empty values in required columns, conversion of variables to a consistent data type, and removal of duplicated records. The cleaned datasets are then stored and exported as standardized files (e.g., ATC_cleaned.csv and CPRD_cleaned.csv) for use in subsequent steps. Both the original and the cleaned versions are available for visualization and review through the graphical interface.

### ATC-based codelist generation

The second module supports the construction of medicinal product codelists based on the hierarchical structure of the ATC classification system. ATC codes are organized using string prefixes corresponding to different hierarchical levels, including anatomical main groups (first character), therapeutic subgroups (first three characters), and pharmacological subgroups (first four characters).

Users select groups at the therapeutic and pharmacological levels and specify whether selected groups and their descendants’ codes should be included. The application retrieves matching codes using prefix-based matching, where all ATC codes starting with the selected prefix are retrieved when descendants’ inclusion is specified. When descendant inclusion is not specified, only codes corresponding exactly to the selected level are retained. Generated entries are then added to the working codelist though a button with standardized fields, including coding_system set to “ATC”, and label set to “DC_Proxy”. In addition to selection through the ATC hierarchy, the user can manually make new entries to the working codelist via user input fields, allowing direct specification of codes and associated metadata. For efficiency reasons, duplicate entries are removed based on unique combinations of coding systems and code since different formulations of the same medicinal product will be included.

### Mapping to prodcodes

The third module maps entries from the working codelist to CPRD Aurum prodcodes. Mapping is performed using case-insensitive exact string matching (e.g., pattern-based matching) between medicinal product names in the working codelist and text fields in the CPRD Aurum dataset, including EMIS_term, product_name, and substance_name. Matched records are combined, and duplicate entries are removed based on coding system and code. For each mapped record, the product_name variable is defined as the first non-missing value among product_name, substance_name, and EMIS_term. Mapped entries are assigned “PRODCODEID” as coding_system by default, and user-defined variables (drug_abbreviation and drug_concept) are derived from the working codelist.

An additional classification step assigns each mapped record to a mechanism category (topical vs systemic) using a predefined lookup table located in the Scripts/Dictionaries/ directory created by the lead researcher in this article. Classification is based on string matching against normalized values of product_name, formulation, and administration_route. Matches are evaluated separately for formulation and route-based terms, each associated with a classification label and priority value. When both classifications are available, the assignment of the highest priority (lowest numeric value in the lookup table) is retained. When only one classification is available, it is assigned directly; otherwise, no classification is assigned. An optional filtering step allows restriction of mapped records to as selected mechanism category prior to inclusion in the working codelist.

### Codelist management and export

Entries generated through ATC selection and prodcodes mapping are appended to the working codelist, which can be edited interactively. Users can add, modify, or remove entries.

The final module enables export of the working codelist. Users can preview the dataset, select the output format (csv or Excel file), and specify a file name. The exported file contains the standardized columns of the working codelist. All fields are written as text.

### Software implementation

DrugSet was developed in R 4.4.0 using the Shiny framework. The application uses standard R packages for data manipulation and interface functionality, including shiny, DT, dplyr, and readr.^18–30^ The application is designed to run locally within an R environment. The app is available in Github (https://github.com/vjolahoxhaj/DrugSet) and has been archived on Zenodo (DOI: 10.5281/zenodo.21257373).^31^

### Validation and performance assessment

Two validation processes were defined: validation of the ATC-based codelist generation and validation of prodcodes mapping. All runtime and performance assessments of the DrugSet app were conducted on a macOS device with an M3 processor and 8GB of RAM.

Reference codelists were constructed using the WHO ATC/DDD Index (2025 version) for selected therapeutic areas, including metformin (A10BA02) alone and combinations, beta-blocking agents (C07A), and topical salicylic acid (D01AE12).^17^ ATC codes at different hierarchical levels were included to represent varying levels of specificity.

ATC codelists generated by DrugSet (2025 version) were compared with the reference codelists using counts if true positives, false positives, false negatives, and true negatives. Sensitivity, specificity, and positive predictive value were calculated using standard definitions. The complete cleaned ATC codelist was used for calculating true negatives.

Prodcodes mappings generated by DrugSet (December 2023, December 2024 and June 2025 version) were compared with a manually curated reference dataset created independently by C.F., D.E.M., S.L., and T.A. (June 2025 version), hereafter referred to as prodcodes manual reference. Comparisons included identification of overlapping codes and codes unique to each mapping. Agreement was calculated as the proportion of overlapping codes relative to the total number of unique codes across both mappings, and absolute differences in code counts were also recorded. The time required to generate codelists using DrugSet, to construct the reference codelists and manual mapping was recorded.

A sensitivity analysis was conducted to assess the impact of differences in CPRD input files on prodcodes mapping. The mapping procedure was repeated using the modified DrugSet input files supplemented with missing codes identified through the main analysis. These were codes present in the input files used for the manual prodcodes reference but absent from the DrugSet version. Mappings generated using these files were compared with the manually curated reference mapping.

## RESULTS

DrugSet was applied to generate ATC-based and CPRD-based codelists for three therapeutic classes: metformin, beta-blocking agents, and topical salicylic acid. For ATC-based codelists, the number of codes generated by DrugSet was identical to the corresponding WHO reference codelists for all evaluated medicinal product classes. For prodcodes mappings, the number of codes generated by DrugSet was comparable to those obtained from manually curated prodcodes reference. The number of codes generated is summarized in Figure 3.

**Figure 3.**
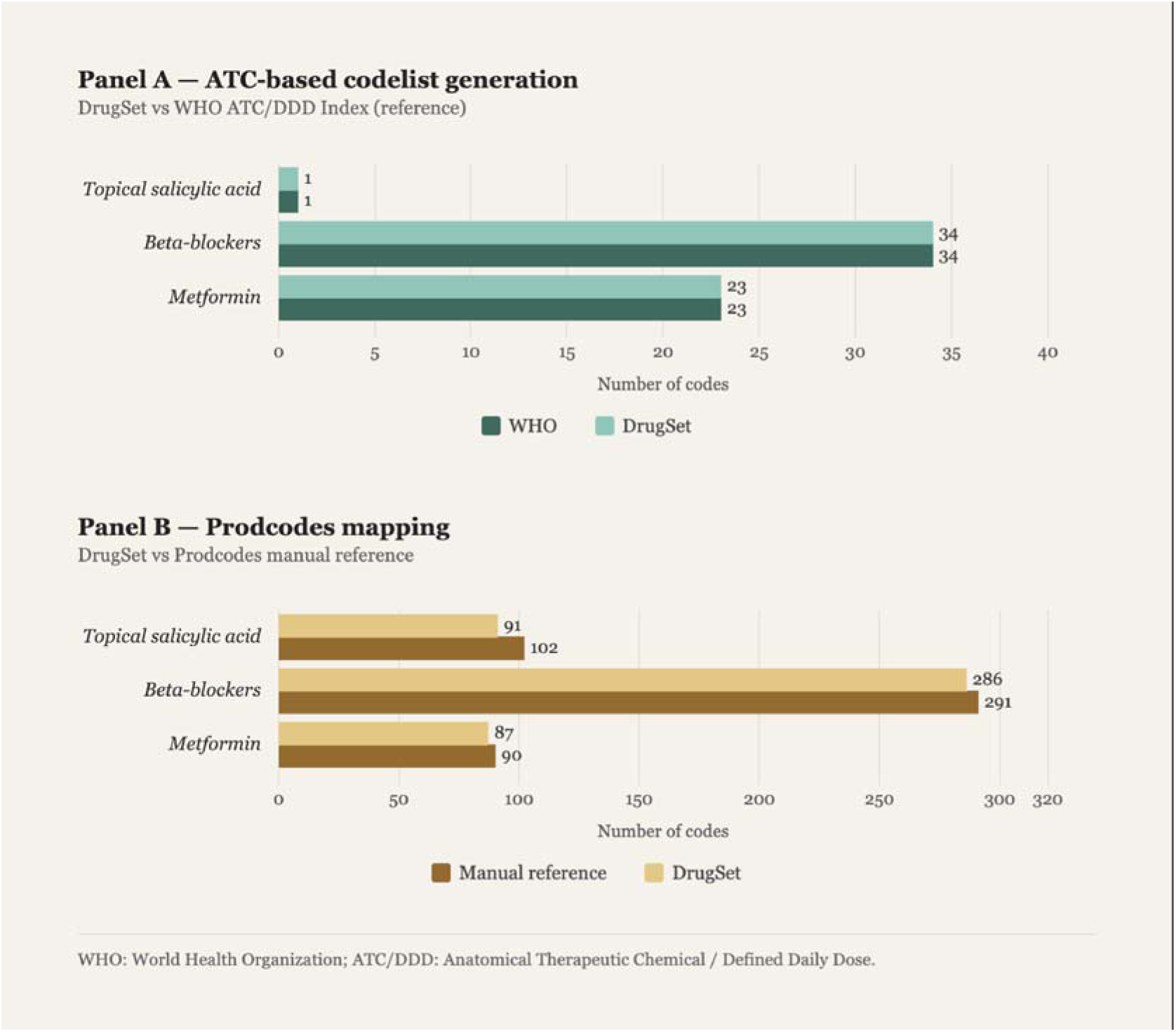
Comparison of medicinal product codelists sizes generated by DrugSet versus reference standards for ATC-based codelists (Panel A) and prodcodes mapping (Panel B). WHO: World Health Organization ATC/DDD: Anatomical Therapeutic Classification/Defined Daily Dose Index.

### Validation of ATC codelist generation

ATC codelists generated by DrugSet were compared with reference codelists derived from the WHO ATC/DDD Index. For all evaluated therapeutic classes, complete agreement was observed between DrugSet-generated and reference codelists. The number of true positives corresponded to the total number of reference codes for each medicinal product class, with no false positives or false negatives identified. The true negatives reflect the remaining ATC codes in the source ATC file (N = 6897) not included in either DrugSet or reference codelists. The metrics for ATC-based codelist validation are presented in Table 1.

**Table 1.** ATC-based codelist generation metrics.

| Medicinal product class | TP | FP | FN | TN | Sensitivity (%) | Specificity (%) | PPV (%) |
| --- | --- | --- | --- | --- | --- | --- | --- |
| Metformin | 23 | 0 | 0 | 6876 | 100 | 100 | 100 |
| Beta-blockers | 34 | 0 | 0 | 6863 | 100 | 100 | 100 |
| Topical salicylic acid | 1 | 0 | 0 | 6896 | 100 | 100 | 100 |

*TP: True positives; FP: False positives; FN: False negatives; TN: True negatives*

### Validation of prodcodes mapping

Prodcodes mapping generated by DrugSet were compared with manually prodcodes reference. Across all evaluated therapeutic classes, high agreement proportions were observed between DrugSet-generated mapping and the reference mappings, ranging from 89.2% - 98.3%. Differences between mappings were characterized by codes unique to either DrugSet or the reference mappings. The metrics for prodcodes-based codelist validation are presented in Table 2

**Table 2.** Prodcodes-based codelists mapping metrics.

| Medicinal product class | Overlapping codes | DrugSet only | Prodcodes reference only | Total unique codes | Agreement (%) |
| --- | --- | --- | --- | --- | --- |
| Metformin | 87 | 0 | 3 | 90 | 96.7 |
| Beta-blockers | 286 | 0 | 5 | 291 | 98.3 |
| Topical salicylic acid | 91 | 0 | 11 | 102 | 89.2 |

The time recorded for generating the final medicinal product codelist using DrugSet ranged from 2 to 4 minutes while the total time using manual mapping (ATC reference codelists and prodcodes mapping) ranged from 27 minutes to 1 hour and 31 minutes. Time diagnostics are presented in Table 3.

**Table 3.** Time diagnostics between manual and automatic codelists creation.

| Medicinal product class | ATC reference | Prodcodes reference | Manual | DrugSet |
| --- | --- | --- | --- | --- |
| Metformin | 6 min | 21 min | 27 min | 4 min |
| Beta-blockers | 7 min | 1 hour 5 min | 1 hour 12 min | 3 min |
| Topical salicylic acid | 3 min | 1 hour 28 min | 1 hour 31 min | 2 min |

### Sensitivity analysis of input data

The sensitivity analysis evaluated the impact of using the modified CPRD Aurum input files on prodcodes mappings. DrugSet mappings generated using the modified-input data were compared with prodcodes reference mappings. The agreement for metformin increased from 96.7% to 100%, and for beta-blockers from 98.3% to 99.6%. The results are summarized in Supplementary table S1.

## DISCUSSION

In this article, we present DrugSet, an R Shiny application designed to support the construction of codelists for medicinal products by integrating ATC-based hierarchical code definition with automated mapping to CPRD Aurum prodcodes within a single workflow. Validation against WHO reference codelists demonstrated 100% sensitivity, specificity, and PPV for ATC codelist generation across three therapeutically and structurally distinct medicinal product classes. Mapping to prodcodes showed high agreement with manually curated reference codelists (89.2%-98.3%), with discrepancies largely due to missing or non-informative fields and absent codes in the CPRD Aurum dictionary used as input. In addition to its accuracy, DrugSet substantially reduced the time required for codelist construction from 3 hours and 10 minutes to 9 minutes across all selected medicinal products.

### DrugSet in the context of existing tools

The development of harmonized codelists for medicinal products in pharmacoepidemiologic research has received increasing attention, and several tools have been developed that address different aspects of this challenge.

OpenCodelists, developed as part of OpenSAFELY platform, provides a collaborative web interface for building, versioning, and sharing codelists primarily based on dm+d and SNOMED terminologies used in NHS data.^13,14,32^ However, this tool is designed primarily for NHS-specific vocabularies and does not natively support the ATC classification system or provide an integrated mapping workflow for ATC to prodcodes. In contrast, DrugSet is built around the ATC/CPRD Aurum axis, addressing the workflow needs of mapping prodcodes starting from a standardized ATC codelist.

Within the OMOP ecosystem, CodelistGenerator (using ATHENA in the background), developed for the DARWIN EU network, supports systematic retrieval of codes for medicinal products from standardized OMOP dictionaries.^16,33^ The DARWIN EU phenotyping network, which incorporates CodelistGenerator alongside a 14-step standard operating procedure, provides a structured reviewed approach to codelist generation.^34^ However, these tools require that data have already been converted to the OMOP Common Data Model (CDM), a process that for medicinal product data involves upfront mapping of source dictionaries to RxNorm, the standard medicinal product dictionary in OMOP. RxNorm is not used in European data sources, and this mapping introduces an additional layer of potential misclassification. In contrast, development of medicinal product codelists using DrugSet does not rely on a CDM-specific dictionary but operates directly on native dictionary files.

Graul et al. described a standardized, reproducible methodology for the construction of codelists for medicinal products in CPRD Aurum, which provides important guidance on handling missing data in product dictionaries such as substance name, formulation, and route of administration.^8^ Their framework emphasizes structured searching across multiple text fields and has been cited as a methodological reference point for CPRD database users. DrugSet is directly aligned with these principles: it searches across the EMIS term, product name, and substance name fields, applies mechanism-based classification using formulation and route of administration fields, and handles missing data through a priority-based assignment logic.

More recently, CodeMergeR, a Shiny-based application for standardization, cleaning, and integration of codelists in the Vaccine Monitoring Collaboration for Europe (VAC4EU) research network, demonstrated the value of automated codelist harmonization in multi-database studies.^35^ While CodeMergeR operates downstream of codelist creation (merging and validating independently produced lists), DrugSet operates upstream, at the point of codelist generation itself. These tools address complementary problems: DrugSet creates and validates individual drug codelists, while CodeMergeR assembles codelists from multiple contributors into a study-ready master file.

A notable development in the field has been the application of large language models (LLMs) for drug classification. Ogorek et al. used GPT-4o with chain-of-thought prompting (i.e., the model is instructed to reason through the drugs mechanism and dose before outputting an answer) to classify free-text daily dose strings, such as those found in unstructured RWD, into second-level ATC codes, demonstrating that the approach could handle heterogenous input formats.^36^ However, the study used only a single medicinal product (aspirin) and evaluated only 200 records, leaving performance across different medicinal product classes unknown. The data was retrieved from a specific population (i.e., patients using a proprietary smart dispenser), and the findings cannot be generalized to EHR, claims or primary care data. Moreover, LLM outputs are probabilistic, and the same input may yield different classification across runs, model versions, or prompt formulations. In addition, prompts themselves are not always openly available, limiting reproducibility. These characteristics make LLM-based approaches difficult to audit and not suitable as the basis for a shareable, version-controlled codelist.

DrugSet is positioned within a growing environment ecosystem of codelists tools, distinguished by its focus on the ATC-CPRD Aurum mapping workflow, its operation on native dictionaries, its mechanism-based classification, and its integration of codelist generation and export within a single interactive environment. To our knowledge, no existing open-source tool provides this specific combination of functionality.

### Strengths and limitations

DrugSet offers several strengths. First, it integrates multiple steps of the medicinal product codelists construction workflow into a single interactive application. This reduces the risk of errors that arise when the process is preformed separately using disparate scripts or manual workflows such as copying and pasting and is consistent with recommendations for reproducible codelist development.^37^ Second, DrugSet achieved perfect agreement with WHO reference codelists (ATC-based codelists) and high agreement with manually curated prodcodes mappings. The discrepancies observed in prodcodes mappings were systematically characterized and were attributable to data quality issues in the input files, rather than errors in the DrugSet algorithm as demonstrated by the sensitivity analysis. Third, the time savings associated with DrugSet are substantial; the application completed codelist construction in 2 to 4 min for all medicinal product classes, compared to 27 min to over 1.5 hours using manual approaches. This gain is particularly relevant in multi-drug studies, where codelist construction for many medicinal product classes can represent a significant proportion of the study’s preparation time. Fifth, the application supports a mechanism-based classification step using a configurable lookup table, enabling researchers to restrict mappings to specific routes of administration or formulations. Sixth, DrugSet allows for multiple CPRD input files and combines them into a single master input file before mapping. This enables users to include several CPRD dictionary releases simultaneously and reduces dependence on a single dictionary version.

Several limitations of this tool should be acknowledged. First, the validation was conducted on three medicinal product classes that were selected to represent varying levels of specificity and complexity. While results were consistent across these classes the generalizability of the validation findings to more complex medicinal products (e.g., combination therapies or drugs with multiple therapeutic indications) cannot be assumed. In addition, DrugSet has not yet been tested for the creation of the vaccines codelists, however given that the process of identifying and selecting vaccine concepts is very similar to medicinal products we expect the same results, but further testing is warranted. Second, DrugSet relies on the completeness and accuracy of the source dictionary files provided as input. Although this is a property of the underlying data rather than the algorithm, users must ensure that the input dictionary files correspond to the version of the data they need. This issue is not native to DrugSet and reflects broader issues of dictionary versioning in research. Because CPRD dictionaries are updated periodically, future versions of DrugSet could include the functionality to record the specific dictionary release used for each mapping. Third, the mapping approach is based on string matching and while this is consistent with established CPRD codelist methodology, it may not capture all relevant codes if relevant information is recorded in non-standard fields (i.e., other variables of the CPRD dictionary).^8^ Fourth, although DrugSet improves the efficiency and reproducibility of medicinal product codelist construction, the generated codelists should not be considered a replacement for clinical or pharmacological review. Expert review remains necessary to confirm whether the mapped products are appropriate for the intended study question, particularly for combination therapies or when the route of administration or formulation are important. Finally, the current version of DrugSet does not support integration with OMOP CDM dictionaries or other non-CPRD Aurum data sources such as dm+d, RxNorm or the EMA’s IDMP. This limits its direct applicability to research networks using other data sources with different dictionaries.

DrugSet supports several key principles that are increasingly advocated in pharmacoepidemiology: the use of structured and hierarchical medicinal product classification, the systematic handling of missing or non-informative data in product dictionaries, mechanism-based filtering, and the production of standardized, exportable codelist files.^8,37^ Adoption of DrugSet within a workflow could further enhance transparency and reproducibility of results by enabling fully traceable, rule-based codelist generation that can be readily audited and reviewed.

## CONCLUSION

DrugSet is a validated, open-source R Shiny application that supports the construction of medicinal product codelists based on the ATC classification system and their mapping to CPRD Aurum prodcodes within a unified, interactive workflow. Validation demonstrated 100% accuracy for ATC codelist generation and high agreement with manually curated prodcodes mappings. The application substantially reduced the time required for codelist construction compared to manual approaches.

## AUTHORS CONTRIBUTION

V.H.: Conceptualization, Methodology, Investigation, Software, Writing – Original Draft. C.L.A.N. and M.C.J.M.: Supervision, Writing – Review & Editing. D.E.M., S.L., C.F., S.D and T.A.: Investigation, Validation, Writing – Review & Editing.

## Data Availability

All data produced in the present work are contained in the manuscript.

## ACKNOWLEDGMENTS

We would like to acknowledge Dr. Sandeep Dhanda for reviewing the prodcodes manual reference codelists used in this study and Drs. Sima Mohammadi for providing the CPRD input dictionary for the DrugSet app.

## SUPPLEMENTARY MATERIAL

**Supplementary file 1. Sensitivity analysis: Prodcodes manual reference vs DrugSet mappings (modified input)**

## DRUGSET AVAILABILITY

DrugSet is available as open-source R shiny app with an MIT Non-Commercial License in the following link https://github.com/vjolahoxhaj/DrugSet.

## DECLARATION OF GENERATIVE AI IN SCIENTIFIC WRITING

During the preparation of this work, the authors used GTP-4 to improve readability and language. After using this tool, the authors reviewed and edited the content as needed and take full responsibility for the content.

