## Supplemental File 1 for "DrugSet: A validated R Shiny application for reproducible drug codelist construction from ATC classification to CPRD Aurum prodcodes"

*Table S1. Sensitivity analysis: Prodcodes manual reference vs DrugSet mappings (modified input)*

| Drug class | Overlapping codes | DrugSet only | Prodcodes reference only | Total unique codes | Agreement (%) |
| --- | --- | --- | --- | --- | --- |
| **Metformin** | 90 | 0 | 0 | 90 | 100 |
| **Beta-blockers** | 290 | 0 | 1 | 291 | 99.6 |
